# Occurrence of Four Dengue Virus Serotypes and Chikungunya Virus in Kilombero, Tanzania during Dengue Outbreak in 2018

**DOI:** 10.1101/2020.10.09.20209783

**Authors:** Beatrice Chipwaza, Robert David Sumaye, Maja Weisser, Winfrid Gingo, Nicholas Kim-Wah Yeo, Siti Naqiah Amrun, Fredros O. Okumu, Lisa F.P. Ng

**Affiliations:** St. Francis University College of Health and Allied Sciences (SFUCHAS), P.O. Box 175, Ifakara, Tanzania; Environmental Health and Ecological Sciences Department, Ifakara Health Institute, P. O. Box 53, Ifakara, Tanzania; Division of Infectious Diseases & Hospital Epidemiology, University Hospital Basel; St. Francis Referral Hospital (SFRH), P.0. Box 73, Ifakara, Tanzania; Infectious Diseases Horizontal Technology Centre (ID HTC), Agency for Science, Technology and Research (A*STAR), Immunos, Biopolis, Singapore 138648, Singapore; Singapore Immunology Network (SIgN), Agency for Science, Technology and Research (A*STAR), Immunos, Biopolis, Singapore 138648, Singapore; Faculty of Health Science, School of Public Health, University of the Witwatersrand, Johannesburg, South Africa; School of Life Science and Biotechnology, Nelson Mandela African Institution of Science and Technology, P. O. Box 447, Arusha, Tanzania; Institute of Biodiversity, Animal Health and Comparative Medicine, University of Glasgow, Glasgow, G128QQ, UK; Department of Biochemistry, Yong Loo Lin School of Medicine, National University of Singapore, Singapore, Singapore; National Institute of Health Research, Health Protection Research Unit in Emerging and Zoonotic Infections, University of Liverpool, Liverpool, United Kingdom; Institute of Infection, Veterinary and Ecological Sciences, University of Liverpool, Liverpool, United Kingdom

**Keywords:** Dengue virus, dengue serotypes, chikungunya virus, Kilombero district, Tanzania

## Abstract

**Background:** Dengue and Chikungunya viruses can cause large-scale epidemics with attack rates exceeding 80%. In Tanzania, there have been repeated outbreaks of dengue fever, the most recent one in 2018 and 2019 mostly reported in coastal areas. Despite its importance, there is limited knowledge on epidemiology of dengue (DENV) and chikungunya (CHIKV) in Tanzania. This study was conducted to investigate the prevalence of DENV and CHIKV in Kilombero district, South-Eastern Tanzania.

**Methods:** A cross-sectional study was conducted at Kibaoni Health Center, in Kilombero district, in the rainy and dry seasons of 2018. Febrile patients of any age and gender were enrolled. Blood samples were taken and screened for DENV and CHIKV viral RNA by real-time RT-PCR assays.

**Results:** A total of 294 patients were recruited. Most were females (65%), and aged between 14⍰25 years (33%). DENV and CHIKV were detected in 29 (9.9%) and 3 (1.0%) patients, respectively. DENV was detected across all age groups and during both dry and rainy seasons. Although all four DENV serotypes were detected, serotypes 1 and 3 dominated and were present in 14 patients (42.4%) each. Additionally, the study showed DENV-1 and DENV-3 co-infections.

**Conclusion:** This study reveals the co-circulation of all four DENV serotypes and CHIKV in Kilombero district. Importantly, we report the first occurrence of DENV-4 in Tanzania. Unlike previous DENV outbreaks caused by DENV-2, the 2018 outbreak was dominated by DENV-1 and DENV-3. Occurrence of all serotypes suggests the possibility of having severe clinical outcomes in future DENV epidemics in Tanzania.

## Introduction

Dengue (DENV) and Chikungunya (CHIKV) viruses pose a significant public health threat with increasing geographic range and severity. There have been several acute onset large-scale epidemics with attack rates of DENV and CHIKV reaching 90% and 85%, respectively [1-3]. Annually, DENV causes up to 400 million infections [4] while CHIKV has been reported in nearly 40 countries around the world with an estimate of 2 million cases [5]. Numerous outbreaks have been noted in Africa, including in East-African countries such as Kenya, where outbreaks have been reported in 2016, 2019 and 2020 [6, 7]. The clinical manifestations of both diseases are similar to malaria; these include fever, rash, arthralgia, nausea, vomiting, abdominal pain and headache. This has led to misdiagnosis and underreporting of such infections in the absence of specific laboratory diagnostic testing, which remains as a major prevailing challenge for health systems in developing countries [8].

The vector of transmission for both viruses is the day-biting *Aedes aegypti* mosquito, which is commonly found throughout the tropical and subtropical areas [9, 10]. The diverse breeding habitats for *Aedes* mosquitoes account for DENV and CHIKV epidemics during the dry seasons and contribute to urban transmission of the diseases [11-13].

In Africa, out of the four immunogenically and genetically distinct DENV serotypes (DENV 1-4), DENV-2 is the most frequently documented [14-17]. Importantly, exposure to one DENV serotype does not confer immunity to other serotypes [18] and secondary infections by another serotype or mixed infections increase the risk of developing dengue hemorrhagic fever (DHF) and dengue shock syndrome (DSS), both being potentially lethal manifestations of dengue [19]. The three genotypes of CHIKV are Asian, West African and East/Central/South African (ECSA) [20]. Since its discovery in 1952 in Tanzania, there have been several reports from studies mostly conducted in Northern and Central Tanzania [15, 21, 22].

In Tanzania, DENV outbreaks were reported in 2010, 2012-2014 [14, 23-25] and recently in 2018 and 2019 [26]. The 2014 outbreak resulted in 1,018 confirmed cases and 4 deaths mainly in Dar es Salaam [11], and a few confirmed cases in Kilosa district [15]. However, the worst documented dengue outbreak occurred in 2019 in Dar es Salaam and later in Tanga [26] with 6,873 cases and 13 deaths [26]. CHIKV infections on the other hand have been documented mostly in few districts in Northern and Central Tanzania with a prevalence ranging from 4% to 7% [15, 22]. Of these outbreaks, there is little information regarding the prevalence of the circulating DENV serotypes and CHIKV. Unfortunately, there is also lack of appropriate pathogen diagnosis in febrile diseases and no surveillance programs have been conducted so far. Therefore, sporadic cases may have occurred, but may have been undetected or misdiagnosed as malaria or other viral diseases because of similar clinical symptoms. The aim of this study was to determine the prevalence of DENV and CHIKV, including identification of DENV serotypes, in Kilombero district, South-eastern Tanzania.

## Methods

### Study setting

This study was conducted in Ifakara Town Council (ITC) located in Kilombero district, South-eastern Tanzania. The Kilombero district is surrounded by wildlife management areas, rainforests and savannahs. The Kilombero River runs through the valley, creating one of the largest flood plains in Africa. The area has a dry season lasting from June to November, and two rainy seasons with heavy rains in March to May, and short rains in December and January. Administratively, ITC has one division subdivided into nine wards and 11 villages. Five wards are in urban areas while four are in rural areas. This study was conducted at Kibaoni Health Center (KHC) located within ITC. KHC serves a population of about 106,424 people mainly from ITC and other parts of the Kilombero and Ulanga districts.

### Study design and population

A cross-sectional study of patients presenting to the KHC with fever was conducted from March to May 2018 (rainy season) and from June to October 2018 (dry season). Eligible criteria were febrile patients of any age and gender with a measured axillary or rectal temperature of ≥ 37.5°C or ≥38°C, respectively. Patients were excluded from the study if they presented with severe illnesses that required immediate medical care, and individuals with positive malaria results by rapid test were excluded in the subsequent laboratory analysis.

### Sample Size

The sample size was estimated to be 334 and was calculated with the formula reported in Naing et al. (2006), using a prevalence of 29% based from a study conducted in febrile patients in Kilosa district, located 180km from the study area [15, 27]. The assumptions for the confidence level and margin of error were 95% (1.96) and 5% (0.05) respectively, while contingencies such as recording error was 5%.

### Clinical examination and sample collection

A trained physician at KHC obtained informed consent from eligible febrile patients. Information on demographics was collected using a standardized questionnaire. A physical examination was performed for each patient. The provisional clinical diagnosis and patient management was done by the hospital clinical team according to the local standard of care. Venous blood (5 ml from children and adults, or 2-3 ml from infants) was collected aseptically into EDTA vacutainer tubes for viral diagnostics. The blood was immediately transported to the Ifakara Health Institute (IHI) laboratory for phase separation. Plasma fractions were obtained and stored at −80°C until analyzed.

### Laboratory analysis

All enrolled patients were initially screened by malaria rapid diagnostic test (mRDT) against malaria parasites using SD BIOLINE Malaria Ag Pf/Pan (Standard Diagnostics, Inc.) at KHC. Samples whose mRDT results were negative were further analyzed by DENV and CHIKV real time Reverse Transcriptase-PCR (RT-PCR) assays.

### Real time RT-PCRs for detection of DENV and CHIKV

RNA was extracted from plasma samples using the QIAamp Viral RNA Mini Kit (QIAGEN, Hilden, Germany). Real-time RT-PCR kits were used for detection and differentiation of CHIKV and DENV 1-4 serotypes (Infectious Diseases Horizontal Technology Centre, A*STAR). The kits comprised of (i) Mix 1 which had specific oligonucleotide primers and Taqman probes for multiplex detection of DENV-1 and DENV-3, and (ii) Mix 2 with specific oligonucleotide primers and probes for detection of DENV-2, DENV-4 and CHIKV, and (iii) internal control (IC) targeting β-actin protein in each respective mix and positive controls for all four DENV serotypes and CHIKV. The real time RT-PCR for both Mix 1 (DENV1 and DENV3) and Mix 2 (DENV2, DENV4 and CHIKV) was carried out in a total reaction volume of 25 μl. For Mix 1, the final primer concentration (forward and reverse) for DENV-1 and DENV-3 were 0.5 μM and 0.2 μM respectively while probes were at 0.2 μM and 0.1 μM respectively. For Mix 2, the final primer concentration (forward and reverse) were 0.5 μM for DENV-2 and CHIKV and 0.2 μM for DENV-4 and probe concentration were at 0.2 μM for DENV-2 and CHIKV, and 0.1 μM for DENV-4. For the internal controls, we used 0.2 μM for both forward and reverse primers, and 0.15 μM for probe. Thermal cycler conditions were used as follows: reverse transcription at 50°C for 20 minutes, initial denaturation at 95°C for 2 minutes and 45 cycles of repeated denaturation at 95°C for 45 seconds and extension at 60°C for 1 minute 15 seconds. One step real time RT-PCR was performed in a Rotor Gene® Q thermocycler (Qiagen).

## Results

A total of 294 febrile patients were recruited in this study. One third, (33.0%) were aged between 14⍰25 years, and another third (30%) were aged 26⍰45 years (Table 1). Most participants were females (65.0%) and half of them (51.7%) were recruited during the dry season. 184 patients (62.6%) presented at KHC with a fever duration of 1-4 days and 101 (34.4%) had duration of 5-7 days. About half of the participants were subsistence farmers (52%) while 12.6% were engaging in business. The rest of the participants were students and children. Administratively, the majority of patients (88.1%) who visited KHC were from ITC and very few came from Kilombero and Ulanga districts. The main presenting symptoms were headache 136 (46.3%) followed by coughing 131 (44.6%), malaise 66 (22.4%), diarrhea 42 (14.3%) and vomiting 9 (0.3%).

**Table 1:**
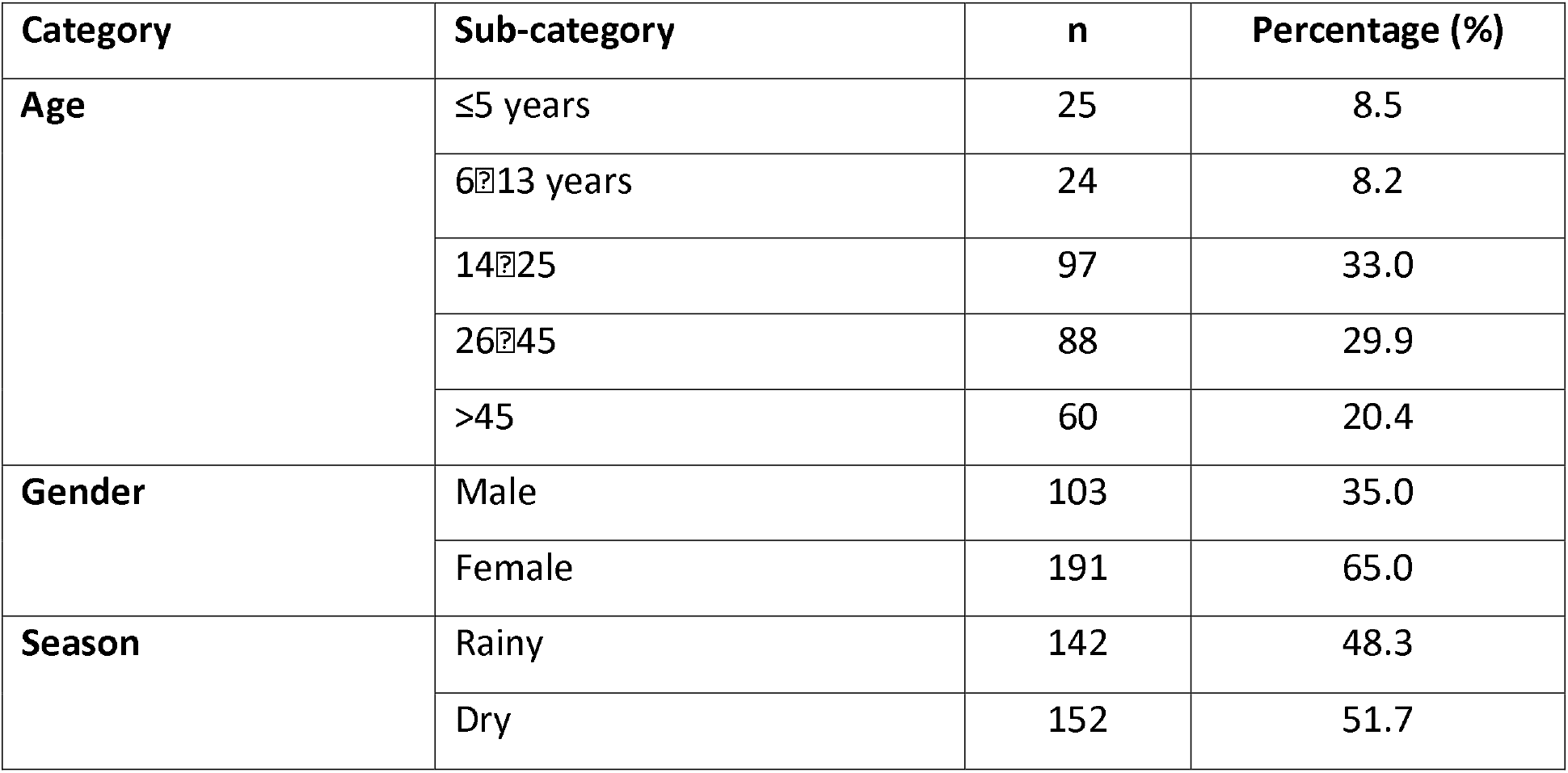
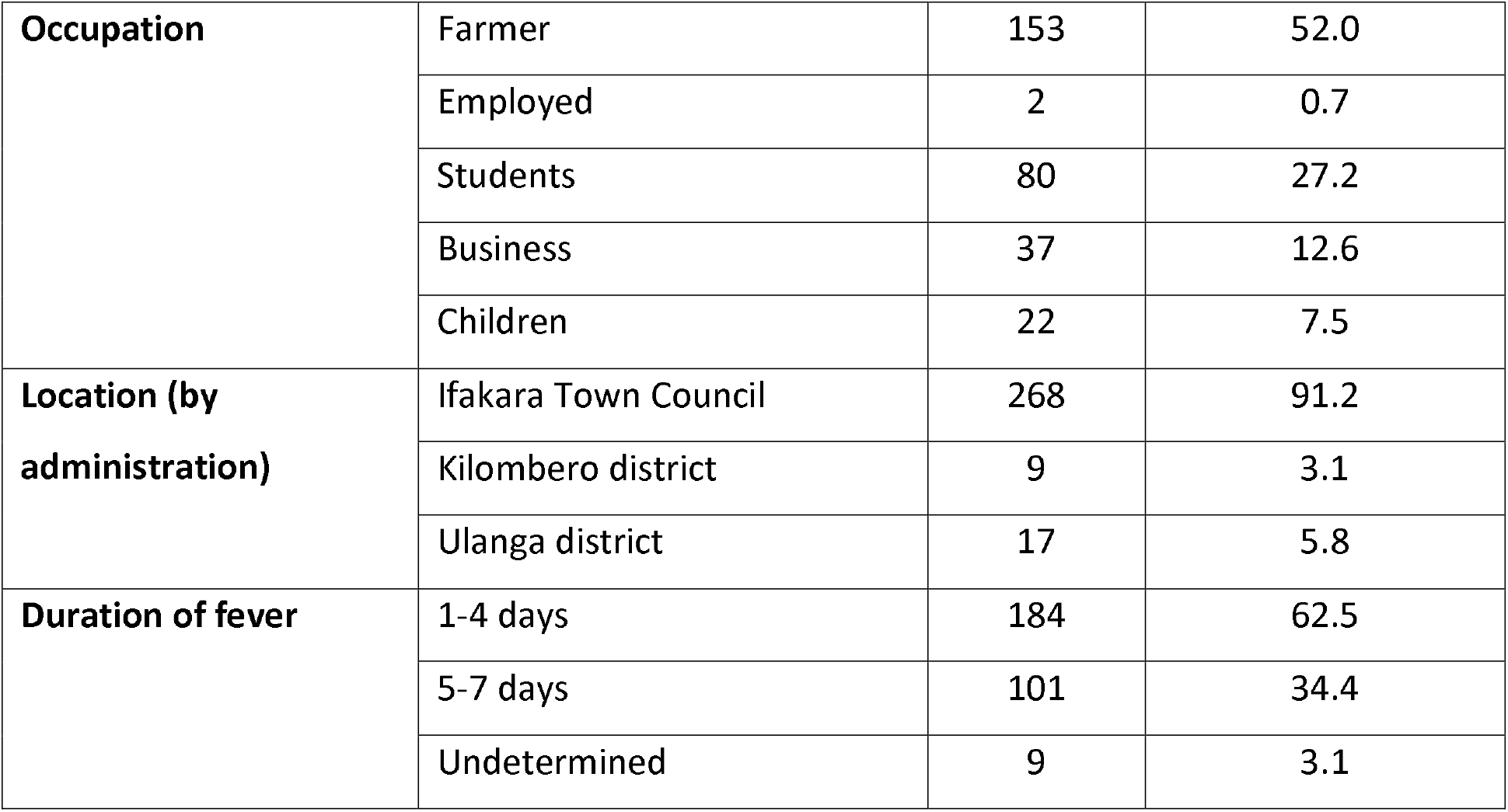
Demographic information of the study participants.

### Prevalence of DENV and CHIKV

Of 294 febrile patients, 29 (9.9%) tested positive for DENV by real time RT-PCR. DENV was detected across all age groups, although the majority was observed in participants aged between 14⍰25 years and > 45 years. Among 29 DENV-positive samples, 19 were females and 10 were males (Table 2). DENV was detected in both seasons, i.e. 16 and 13 cases in rainy and dry seasons, respectively. DENV viral RNA copies were detected mostly in patients with fever duration of 1 – 4 days. In terms of location (by administration), most of the DENV patients came from ITC. In this study, all DENV serotypes were detected but DENV-1 and DENV-3 were more frequent (14 cases each), as summarized in Table 3. DENV-2 was detected in two patients while DENV-4 was observed in one patient. Furthermore, our findings revealed co-infection between DENV-1 and DENV-3 in two patients as indicated in Table 3.

**Table 2:** Prevalence of DENV by real time RT-PCR.

| Category | Sub-category | n (%) |
| --- | --- | --- |
| **Overall** | All patients | 29 (9.9) |
| **Age** | ≤5 years | 3 (1.0) |
|  | 6–13 years | 1 (0.3) |
|  | 14–25 | 10 (3.4) |
|  | 26–45 | 6 (2.0) |
|  | >45 | 9 (3.1) |
| **Gender** | Male | 10 (3.4) |
|  | Female | 19 (6.5) |
| **Season** | Rainy | 16 (5.4) |
|  | Dry | 13 (4.4) |
| **Location** | Ifakara Town Council | 26 (8.8) |
|  | Kilombero district | 2 (0.7) |
|  | Ulanga district | 1 (0.3) |
| **Duration of fever** | 1-4 days | 27 (9.2) |
|  | 5-7 days | 2 (0.7) |

**Table 3:**
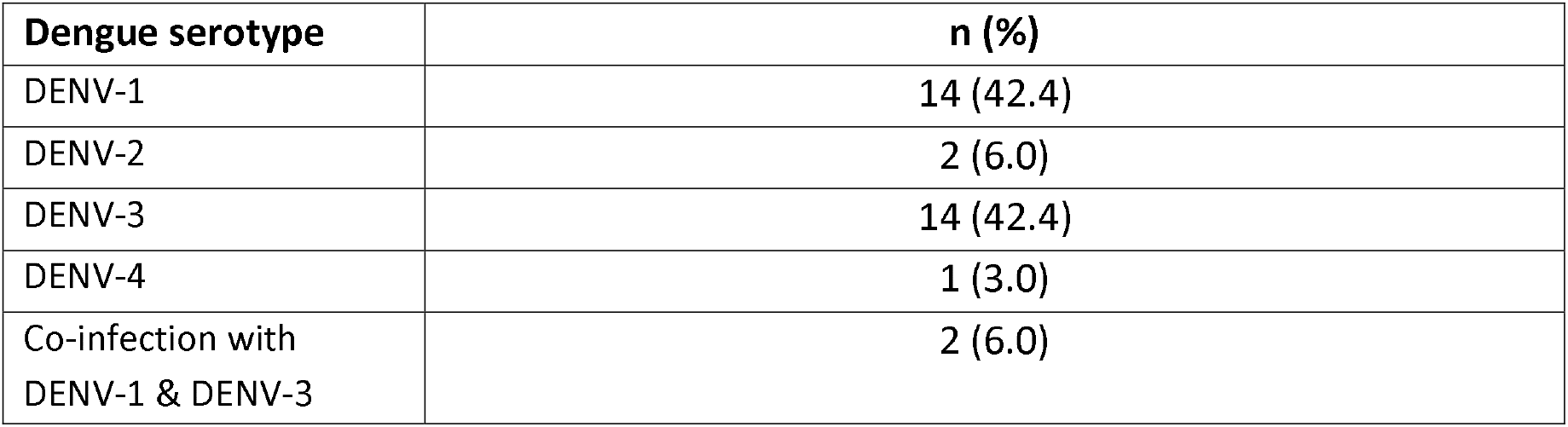
Proportion of DENV serotypes among PCR-positive samples.

Concerning CHIKV, only 3 patients (1%) out of 294 were CHIKV RT-PCR positive. Among the three positives, two were males. The three patients were a 22-year-old student, a 27-year-old farmer, and a 70-year-old farmer. Two CHIKV-positive cases were detected during rainy season and one in dry season.

## Discussion

This study reveals the presence of DENV and CHIKV among febrile patients whose blood samples were collected in 2018 during a DENV outbreak in the coastal area of the country. Our findings have demonstrated the occurrence of all four DENV serotypes as well as a co-infection with DENV-1 and DENV-3.

In the present study, the 9.9% prevalence of DENV was lower than previously reported in Dar es Salaam in 2019 where 17 out of 20 [28]. The difference could be due to the study population, as the study from Dar es Salaam only involved dengue-suspected cases while the current study involved outpatient febrile patients from a a health center. Also, the lower prevalence of DENV in our study could be a result of viral RNA detection sensitivity by RT-PCR declining with duration of fever, and therefore we might have missed patients with >5 days fever duration.

The 1% prevalence of CHIKV observed in this study is lower than that of Northern and Central Tanzania, where previous epidemiological studies reported a prevalence of 4 ⍰ 7% [21, 22]. Furthermore, the low prevalence of CHIKV in Tanzania is in contrast to other countries in Africa (including East Africa) where outbreaks of CHIKV have been frequently reported since 1952 with prevalence up to 53%, n=100/189, [29-32]. Our findings document the persistent circulation of CHIKV, supporting seroprevalence studies in other districts of Tanzania where anti-CHIKV antibodies (both IgM and IgG) were detected i.e. 4.7% (n=364) and 14% (n=105) respectively in the population, thus suggesting the endemicity of the disease [15, 33].

Unlike in the previous DENV outbreaks in Tanzania, which were mainly caused by DENV-2 [15, 25, 34], findings from the current study have shown that all DENV serotypes were circulating, with a predominance of DENV-1 and DENV-3 in the 2018 outbreak. Our results are in line with the findings from the study done in Dar es Salaam in 2019, which reported the co-circulation of DENV-1 and 3 [28]. Furthermore, DENV-1 was detected in a traveler returning from Tanzania to Japan in 2019 [35] and DENV-3 had been reported in Japan from a traveler who had visited Tanzania in 2010 [36], hence supporting its existence in the country. During the repeated DENV outbreaks in Tanzania, the reported disease severity was low to moderate with few deaths, i.e. 4 and 13 deaths in 2014 and 2018-2019 outbreaks, respectively [11]. Out of the four deceased cases in 2014, one had presented with DHF, and one with multiple organ failure. Unfortunately, no data on the DENV serotype is available from those cases [37]. With the increasing number of cases reported recently and the co-existence of all DENV serotypes, more severe disease or fatal outcomes might be observed in the future [19, 38].

The present study was conducted at a healthcare facility serving urban and semi-urban communities. Most DENV-positive cases seen in this study (26 samples) were from ITC and occurred in both rainy and dry seasons. This is in line with other studies, which have demonstrated the occurrence of DENV in urban and semi-urban areas, correlating to the distribution of the vector *A. aegypti* mosquitoes [39, 40]. The surrounding wetlands in our study settings provide suitable breeding habitats for mosquitoes, and in addition the presence of short rains could further favor breeding of *A. aegypti* throughout the year. Thus, this increases the rate of CHIKV and DENV transmission.

### Strengths and Limitations of the Study

To the best of our understanding, this study is the first to show the occurrence of DENV and CHIKV in South-Western Tanzania. The study was conducted at a primary healthcare facility where most patients seek treatment and thus gives a good representation of the general population. However, we tested the samples by real time RT-PCR only, which might lead to under detection of cases with a symptom duration of more than five days. Our findings could be complemented by employing serological tests coupled with acute and convalescent serum, and isolating RNA from whole blood and urine as viral RNA persist longer in the urine.

## Conclusion

This study has demonstrated the occurrence of all four DENV serotypes and CHIKV in Kilombero district, with DENV-4 being reported for the first time in Tanzania. In addition, we revealed the presence of co-infection with DENV-1 and 3 in one patient, possibly corresponding to a changing pattern of DENV serotypes from DENV-2 in the previous outbreaks to DENV-1 and DENV-3 in the 2018 outbreak. These findings are important in planning for future epidemics. We recommend further studies particularly in the identification of local vectors and understanding local transmission dynamics.

## Data Availability

The datasets used and/or analyzed during the current study are available from the corresponding author on reasonable request.

## List of abbreviations

DENV: dengue virus
CHIK: chikungunya virus
RT-PCR: reverse transcription polymerase chain reaction
DHF: dengue hemorrhagic fever
DSS: dengue shock syndrome
ITC: Ifakara Town Council
KHC: Kibaoni Health Center
IHI: Ifakara Health Institute
NTC: Non template control

## Declarations

### Ethics approval and consent to participate

This study was approved by Institutional Review Board of Ifakara Health Institute (IHI/IRB/No: 12-2017) and Medical Research Coordinating Committee of Tanzania’s National Institute for Medical Research (NIMR/HQ/R.8a/Vol.1X/2565). A written informed consent was obtained from each participant. For children under 12 years, a written informed consent was sought from a parent or guardian. In addition, verbal assent was also obtained from children aged 7–12 years, whereas those aged 12 – 18 years provided their own written assent in addition to consent of a parent or guardian. Each patient was assigned a study identification number to ensure confidentiality, which were applied to all patient specimens and data. All data were stored on a password-locked computer.

### Consent for publication

All participants gave consent for publication.

## Authors’ contributions

BC, MW, FO, and LFPN conceived the study; BC, RDS and WG participated in study implementation; BC, NKWY and SNA assisted in laboratory analysis; RDS analyzed the data; BC and RDS prepared the first draft; LFPN contributed reagents. All authors have read and approved the final manuscript.

## Acknowledgements

We would like to thank the Town Council Office of Ifakara and Kibaoni Health Center (KHC) for giving us permission and allowing us to use their facility to conduct this study. We are also grateful to participants for their willingness to participate in this study. We sincerely acknowledge the medical officer in charge of KHC for his cooperation throughout the study period. Special thanks go to the clinicians, Pilly Azizi Ally and Joyce Karlo Mbata, of KHC, and laboratory technician, Sebastian Cobero, of Ifakara Health Institute. Finally, we acknowledge the Infectious Diseases Horizontal Technology Centre (ID HTC) and Singapore Immunology Network (SIgN), Laboratory of Microbial Immunity for providing us multiplex real-time RT-PCR kits for detection and differentiation of CHIKV and DENV serotypes 1-4.

