## Supplementary figures and images for "Occurrence of Four Dengue Virus Serotypes and Chikungunya Virus in Kilombero, Tanzania during Dengue Outbreak in 2018"

### Supplementary Figure 1

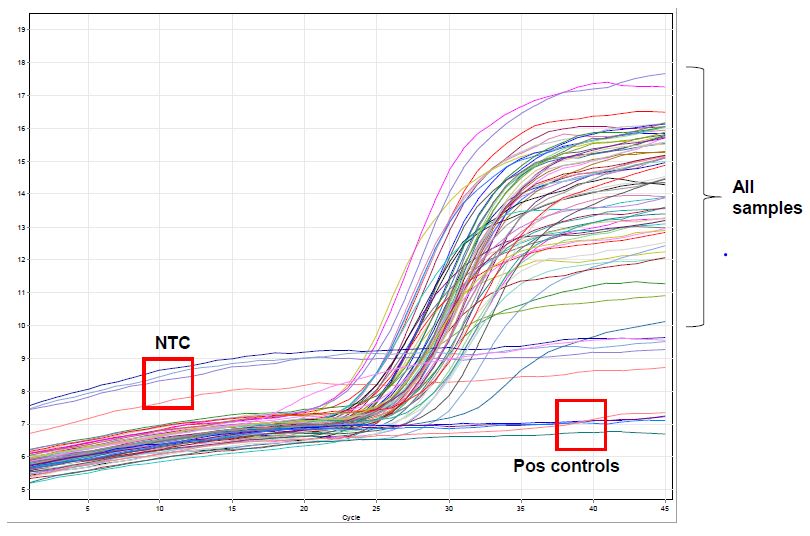

### Supplementary Figure 2

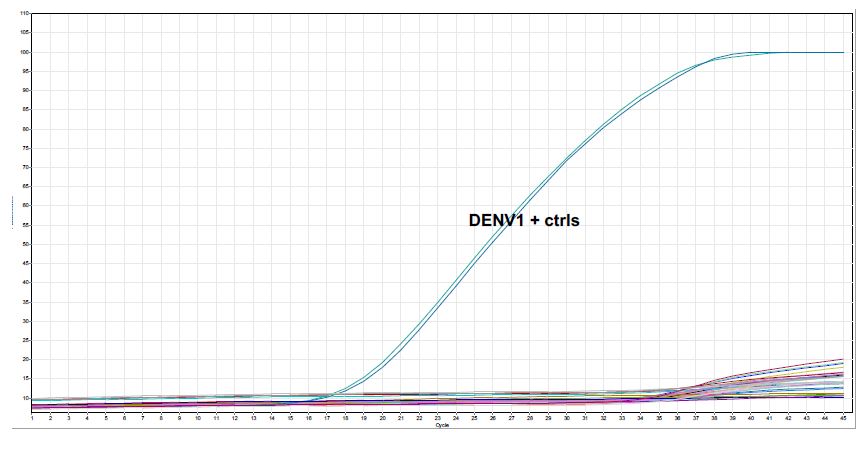

### Supplementary Figure 3

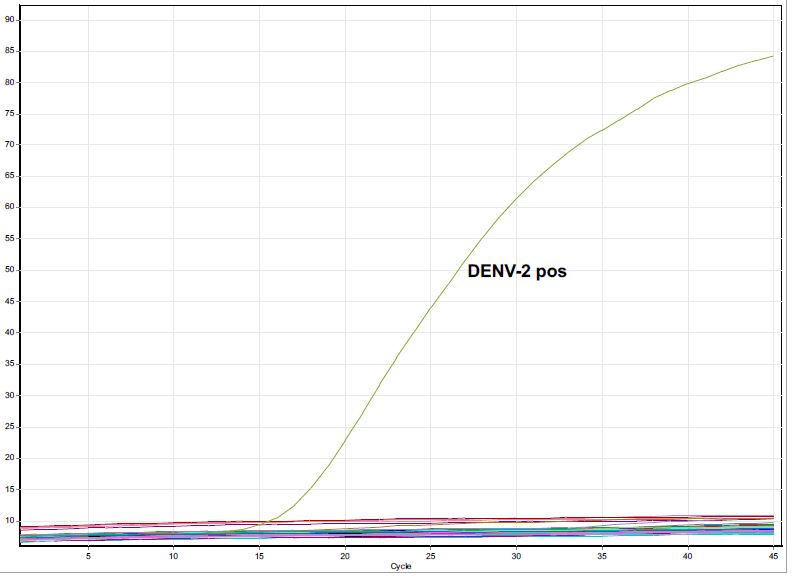

### Supplementary Figure 4

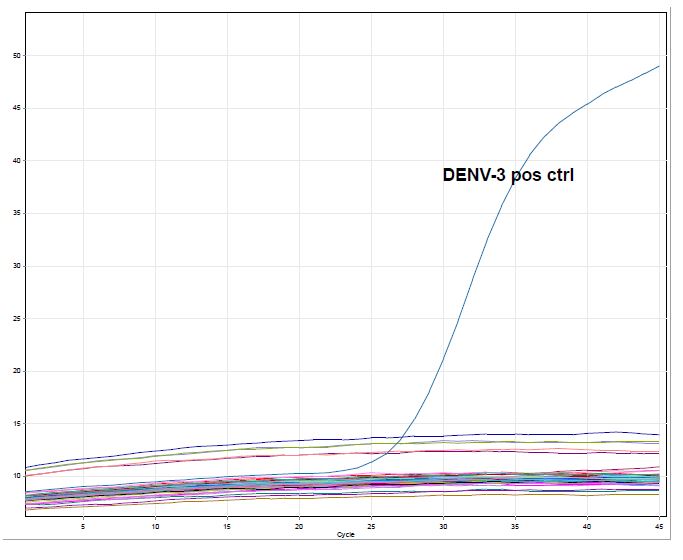

### Supplementary Figure 5

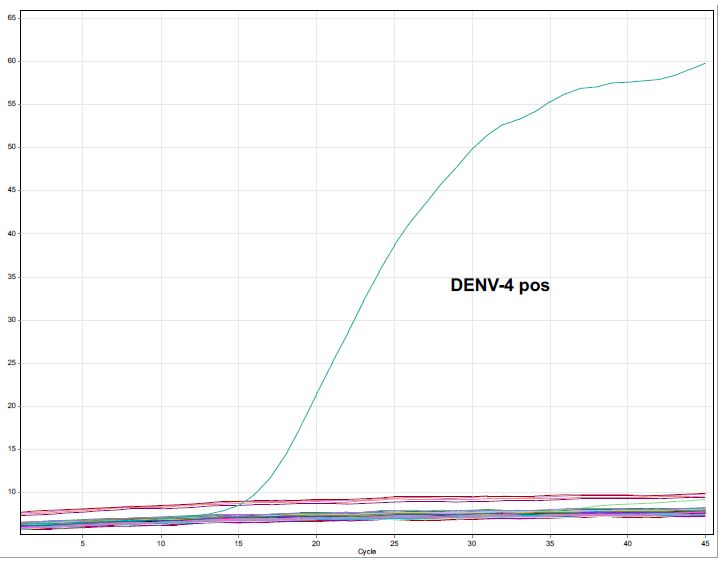

### Supplementary Figure 6

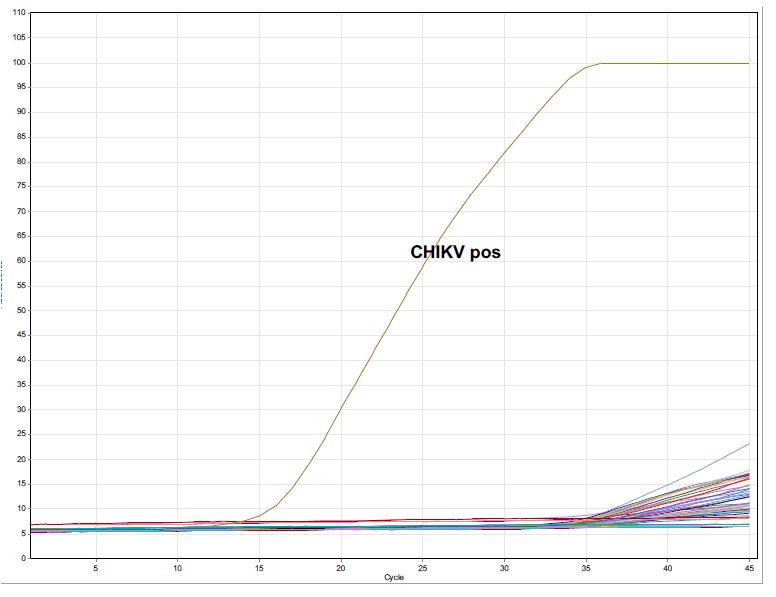
